# Disparities in Naltrexone Prescriptions to Medicaid Enrollees During the COVID-19 Pandemic

**DOI:** 10.1101/2022.10.05.22280706

**Authors:** Hemmy A. Subervi, Lysine E. Varghese, Brian J. Piper

## Abstract

**Background:** The COVID-19 pandemic strained healthcare facilities and the isolation and uncertainty associated with the pandemic compromised mental health around the world. The pandemic has also been associated with an exacerbation of the opioid crisis in the United States (US), and previous studies have reported changing trends in opioid misuse during the pandemic. Our study investigated naltrexone, a prescription drug used to treat alcohol and opioid use disorders by blocking opioid receptors to reduce cravings. We sought to investigate the changes in naltrexone prescriptions issued to Medicaid enrollees in light of the COVID-19 pandemic.

**Methods:** The total number of naltrexone, generic and brand name, prescriptions across the US were obtained from the Medicaid.gov database, expressed as prescriptions per state corrected for the number of enrollees, and organized into two time periods - the pre-pandemic period from January 2019 to December 2019 and the pandemic period from January 2020 to March 2021. Statistical analyses included a paired t-test, a heat map to depict state level variation, and waterfall figures. Procedures were approved by the IRB of Geisinger.

**Results:** There were increases in total naltrexone prescriptions throughout the time frame studied, but a decrease in prescriptions per 100,000 Medicaid enrollees. A paired *t*-test revealed a significant decrease in naltrexone prescriptions during the pandemic period. There was a 398-fold difference between the highest and lowest states in 2019 Quarter 1 and 424-fold in 2021 Quarter 1. Percent change calculations indicated South Dakota (+141%) and Oregon (+172%) showed a significant increase in total naltrexone prescriptions from pre-pandemic to post-pandemic from the national mean (−23.57%±5.60%).

**Conclusions:** The results of this study were significant and indicated a relationship between the COVID-19 pandemic and declining naltrexone prescription rates. Naltrexone prescriptions per 100,000 enrollees decreased in most states during the pandemic and fell by over 32% nationally from 2019 to 2021 despite a slight increase in total prescription numbers and an increase in Medicaid enrollees. These data suggest wide variation in access to substance use disorder treatment during the pandemic. Further research with privately insured patients may be beneficial.

## Introduction

Substance misuse in the United States (US) has been worsening over the past several years and has recently been exacerbated by the COVID-19 pandemic (1). From 2019 to 2020, through the initial stages of the pandemic, overdose death rates increased by 30% (2). This may be attributed to fewer opportunities for distraction from substance cravings, greater ease of access to substances and increased opportunity, loss of social support, and decreased access to treatment (1). The pandemic also elicited a response from the US government to expand Medicaid flexibility and eligibility to help alleviate the unprecedented socioeconomic and psychological stressors that came with lockdowns and isolation (3). Despite the expansion of Medicaid to the public and the increased accessibility of opioid drug treatment across the country, over 70% of drug overdose deaths in 2019 were due to opioids (4). A 2020 study highlighted the increase in alcohol consumption by adults during the COVID-19 pandemic, including a significant 41% increase in the average number of days of heavy drinking (5). Another report determined that despite this increase in use, patients suffering from alcohol use disorder (AUD) were less able to seek treatment (6). Alcohol-related deaths have increased at a rate of roughly 2.2% per year for the past twenty years but increased 25.5% between 2019 and 2020 (7). Despite the growth in availability of substance use disorder medications (8), people who suffer from OUD and AUD are subjected to health care disparities, geographic disparities, and stigmas that surround these diseases (9). Medicaid expansion contributed to a favorable change in health and an overall improvement in self-reported health status among a low-income group (10). This data shows that the increased accessibility of substance misuse medications has a positive impact on communities that are suffering the most.

Naltrexone is a prescription drug that is commonly used to treat OUD and AUD. It acts as an opioid-receptor partial inverse antagonist that decreases cravings associated with substance use disorders (11). Initiation of naltrexone therapy is recommended after symptoms and signs of withdrawal have subsided and at least three days of abstinence from substances (12). The number needed to treat (NNT) to prevent return to drinking was 20 for oral naltrexone and the NNT to prevent return to heavy drinking was 12 (13). Naltrexone can cause both neuropsychiatric and gastrointestinal adverse effects, which have been shown to affect compliance (14). Naltrexone is contraindicated in patients experiencing acute hepatitis and liver failure and who are actively using opioids or experiencing opioid withdrawal (12). Medicaid expansion has made substance use disorder treatments such as naltrexone more accessible to Americans with OUD and AUD. Accessibility to naltrexone is especially vital for Medicaid enrollees who are disproportionately affected by OUD and AUD (15). Medicaid beneficiaries are also treated for substance use disorders more frequently than privately insured adults (16). There had already been suggestions in recent years to expand the availability of medications for addiction treatment drugs to continue combatting OUD in the US (17). However, the unanticipated effects associated with the COVID-19 pandemic prompted citizens to call upon the US federal and state governments to expand flexibility and eligibility for Medicaid enrollment (18). Concerns that the COVID-19 pandemic would devastate an already at-risk population prompted recommendations to increase Medicaid eligibility and allow for more enrollment without inadvertently disenrolling any existing beneficiaries (18). With the heightened rate of opioid and alcohol use and the response to demands for Medicaid expansion, there may be differences in patterns of naltrexone prescription since the start of the COVID-19 pandemic. These patterns have not been described in recent research and there is a lack of information regarding the relationship between the pandemic and naltrexone prescriptions. A study done from 2002 to 2017 investigated the impact of Medicaid expansion on pharmacotherapy for substance use, specifically prescriptions such as injectable naltrexone and buprenorphine (19). This research gives a good insight into how a previous Medicaid expansion affected naltrexone prescription rates, but it does not discuss recent years. Further, Medicaid expansion during the COVID-19 pandemic may have affected the rate of naltrexone prescription as well. Quantifying the changes during this time may inform healthcare providers regarding any upward trend in substance misuse during the pandemic and in treatments administered in response (18). Furthermore, it may provide federal officials with information regarding the effects of the pandemic and Medicaid accessibility on treatment of substance use disorders and how this may affect future Medicaid expansion and drug distribution across all states.

The purpose of this research was to measure the impact of the COVID-19 pandemic on naltrexone prescriptions to US Medicaid enrollees. Our primary objective was to identify a relationship between the consequences of the pandemic with changes in naltrexone prescriptions among Medicaid beneficiaries to help fill the gap in this research pertaining specifically to the duration of the COVID-19 pandemic. We also aimed to describe the overall trend in naltrexone prescriptions over the time period studied and compare prescription rates across the US. We hypothesized that the number of naltrexone prescriptions to Medicaid enrollees during the COVID-19 pandemic would be less than the number of prescriptions prior to the pandemic. We predicted that the shutdowns and social distancing guidelines implemented during the pandemic would lead to reduced access to healthcare and therefore fewer naltrexone prescriptions despite an increase in opioid and alcohol use.

## Methods

### Procedures

All Medicaid enrollees who were prescribed generic or brand name naltrexone between January 2019 and March 2021 were eligible for inclusion. All data was collected from the Medicaid.gov database. New variables were created by calculating the ratios of naltrexone prescriptions per 100,000 enrollees in each of the fifty states and Washington D.C. January 2019 through December 2019) was designated as “pre-pandemic” and January 2020 through March 2021) as “pandemic”. Although the first outbreaks of the COVID-19 virus were detected in late 2019, the United States was not affected until 2020 (20). At the time of data collection (February 2022), Medicaid.gov had updated records through the first quarter of 2021. Quarter 1 of each year includes the months of January, February, and March; Quarter 2 includes April, May, and June; Quarter 3 includes July, August, and September; and Quarter 4 includes October, November, and December. All the collected data was organized by quarter and by state.

### Data-analysis

A paired *t*-test compared pre-pandemic and pandemic values by state using GraphPad Prism version 9.3.1 for Windows. This program was also used to create the figures. A waterfall plot was used to portray percent change in naltrexone prescriptions by state between these time periods.

## Results

The total number of naltrexone prescriptions from January 2019 through March 2021 are represented in Figure 1 and shows pronounced state-level disparities with lowest values in the southeast. Ten of the eleven lowest states had not expanded Medicaid. The mean distribution of naltrexone prescriptions per 100,000 Medicaid enrollees across the US in the pre-pandemic and pandemic time periods are compared in Figure 2. A paired *t*-test at a significance level of α = 0.05 with df=50 and 95% confidence interval was performed for this data and revealed a significant decrease with a *p*-value of <0.0001. In addition to nationwide prescription rates, the percent change in naltrexone prescription rates by state was also calculated. The national mean reduction was −23.57%. These changes are displayed in Figure 3 and the results of these calculations reflect the overall decrease portrayed in Figure 2. A considerable decrease in prescriptions per 100,000 enrollees was observed in all but five states, with only two outliers, Oregon and South Dakota, exceeding a 40% increase.

**Figure 1.**
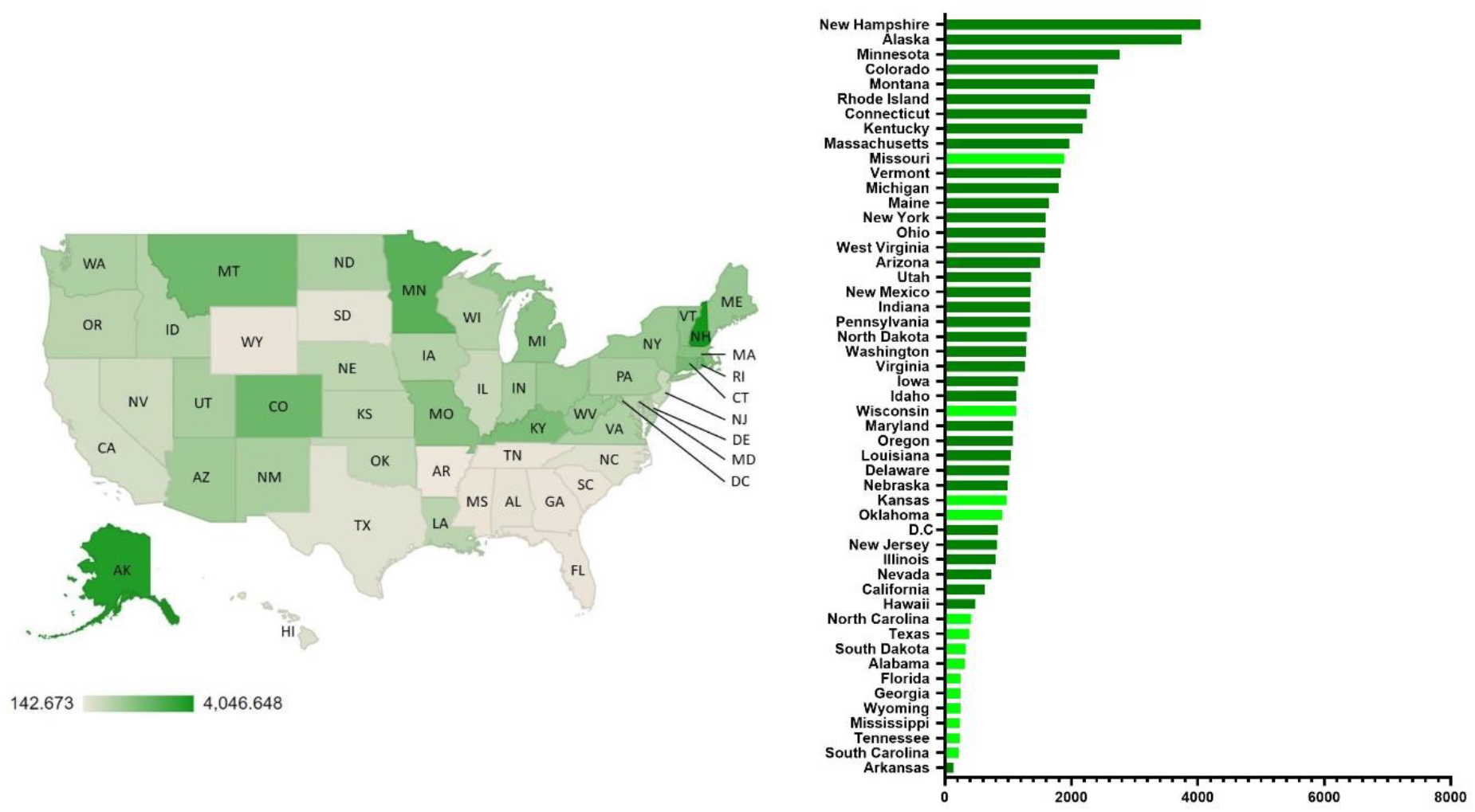
Total naltrexone prescriptions per 100,000 Medicaid patients in the United States from January 2019 - March 2021. On the right, expansion states are shown in dark green and non-expansion states in light green.

**Figure 2.**
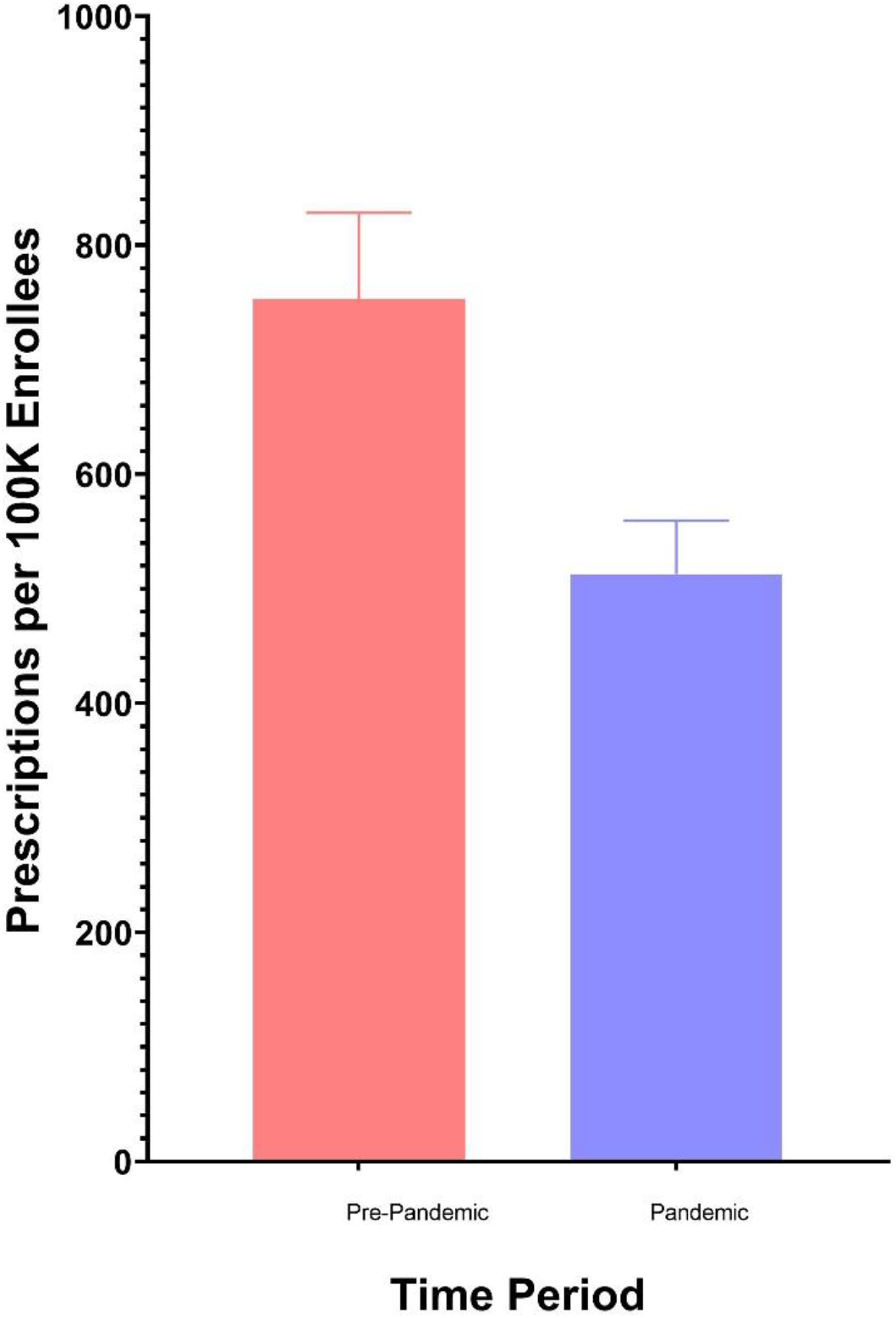
Standard mean distribution of total naltrexone prescriptions per 100,000 Medicaid enrollees for the time-period 2019 (pre-pandemic) and January 2020-March 2021 (pandemic).

**Figure 3.**
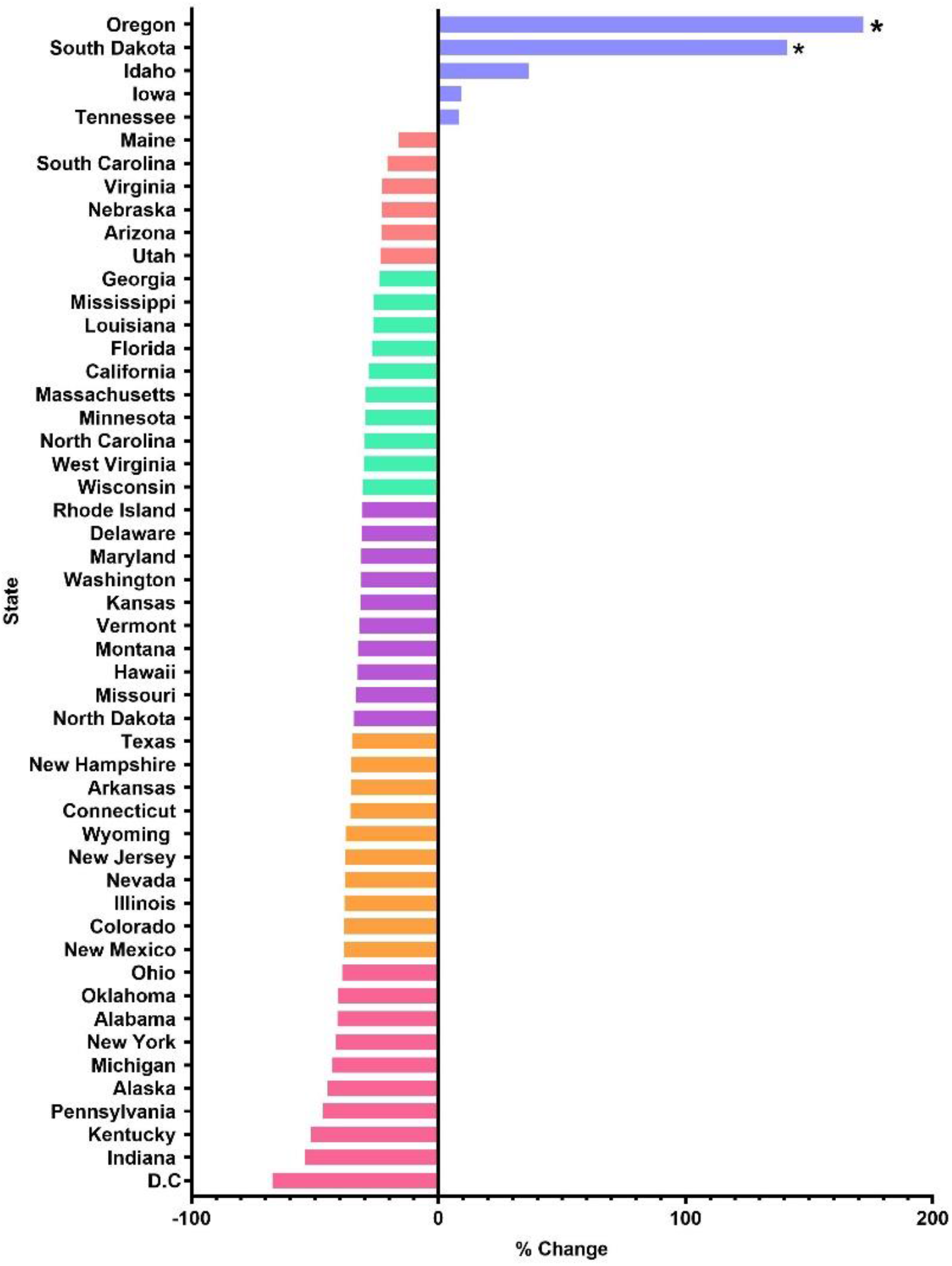
Pandemic and pre-pandemic percent change of total naltrexone prescriptions. Data marked with an asterisk (*) indicates significant difference from the national mean (−23.57% ± 5.603 SEM) from January 2019 - March 2021.

## Discussion

There are quite a few key findings in this data. The results of our study provide additional insight into how naltrexone prescription rates, specifically to Medicaid enrollees, have changed due to the COVID-19 pandemic. The reported *p*-value of <0.0001 indicates that the difference between the prescription rates from the pre-pandemic time period and the pandemic time period were significant. There was an overall decrease in naltrexone prescriptions from the pre-pandemic period to the pandemic period, and almost all states exhibited a negative percent change in prescription rate. Thus, the data is significant for a decrease in naltrexone prescriptions during the COVID-19 pandemic. This supports our initial hypothesis and suggests a relationship between the pandemic and naltrexone prescription rate. Our hypothesis was based on the prediction that the shutdowns and stay-at-home orders put in place during the pandemic would reduce access to healthcare and specifically OUD/AUD treatment. Prescriptions per 100,000 enrollees dropped during the first two quarters of 2020, in the initial stages of the pandemic and lockdown period, and substantially increased afterwards. This could be due to any number of reasons. There was considerable Medicaid expansion in many states during the pandemic and a subsequent increase in total number of enrollees (21), but limited access to healthcare during the pandemic could have contributed to the disparity between Medicaid enrollee increase and naltrexone prescription rate decrease.

The percent change data presented in Figure 3 is also of interest. While the vast majority of states exhibited a considerable decrease in naltrexone prescription rates, just five states experienced an increase. These states were Oregon, South Dakota, Idaho, Iowa, and Tennessee. Of these states that had increases in naltrexone prescription rates, the most notable were the two outliers – Oregon and South Dakota – which both displayed a percent change of over 140% (20). South Dakota is consistently ranked as having very few and lenient COVID-19 restrictions, while Oregon is ranked as having moderate restrictions (22). This discrepancy is interesting because stricter COVID-19 restrictions would likely result in higher rates of unemployment, isolation, and healthcare inaccessibility while more lenient COVID-19 restrictions would likely result in more COVID-19 cases and overwhelmed healthcare facilities, but few limitations on social activity. However, there have been recent drug decriminalization efforts in Oregon, which could contribute to increased substance use and therefore increased need for treatment (23). However, this does not account for South Dakota or imply a causal relationship. Therefore, it is difficult to discern exactly what is causing this dramatic increase in these two states based on this data alone.

The overall data represented in Figure 1 may be of interest for further research. The heat map gives a visual representation of naltrexone prescription disparities across the country. It was expected that states with higher populations, such as California and several states in the northeast, would have the highest rates of prescription. However, the highest rates were seen in Medicaid expansion states that greatly increased their enrollment with new eligibility guidelines. The lowest prescribing states were clustered in the southeast.

It is also worth noting that the long-standing possibility of a vaccine was finally brought to fruition in late 2020, which created a major shift in attitude towards the COVID-19 pandemic (24). This could have resulted in a change in opioid and alcohol use and subsequent changes in naltrexone prescriptions.

A caveat of this report is that naltrexone has several off-label uses. Low-dose naltrexone reduces inflammatory response and can be used as a treatment for autoimmune diseases such as multiple sclerosis and inflammatory bowel disease (25). It can also be used for chronic pain disorders including diabetic neuropathy (25). Considering that the off-label uses of this drug were not specifically investigated in this study, it cannot be concluded that OUD and AUD therapy accounted for all of the naltrexone prescriptions reported between 2019 and 2021. Thus, there could be more complexities in the relationship between increase in naltrexone prescriptions and exacerbation of substance use during the pandemic than suggested by the results of this study.

Including a comparison of the rates of brand name vs. generic naltrexone and rates of the oral formulation vs. injectable naltrexone was considered. However, there was insufficient data in the Medicaid.gov and FDA databases to properly address any differences in these aspects. If any of these data become available in the future, it may be worth considering because of the different purposes for each. For example, AUD is almost always treated with the oral formulation while the injectable may be used for both AUD and OUD.

It is also of note that data was only collected through March of 2021. While the initial impact of the pandemic had mostly subsided by this point, there were still several restrictions in place with the threat of new waves ahead. The mandates and guidelines that intended to slow the spread of COVID-19 were gradually lifted after vaccinations were made available to the public. The first modification of the strict guidelines came in late March of 2021 (19). These fewer restrictions may have changed substance use and healthcare access in the months after the time period discussed here. To more fully assess the effects of the entire pandemic, incorporating data through the end of 2021 would be more representative of the effects of COVID-19. Additionally, previous studies have shown that the COVID-19 pandemic has been associated with an exacerbation of the opioid epidemic in the United States (1). However, it is difficult to ascertain how much of this was directly or indirectly caused by COVID-19. The opioid crisis was likely to have worsened during this period, even in the absence of the pandemic, however the magnitude of increase in overdose deaths from 2019 to 2021 is much higher than would have been otherwise expected (7). Given this uncertainty, it is difficult to conclude that any significant changes in naltrexone prescription patterns to Medicaid enrollees are due to the COVID-19 pandemic or responses to it. Further, due to loss of employment, Medicaid enrollee statuses may have changed during the COVID-19 pandemic which can result in skewed results (26). We accounted for some of this uncertainty by extending the data range to include data from 2019 for comparison. It should be repeated that naltrexone is being studied as a treatment for other disorders as well (16), and these uses have not been considered in this descriptive study. Limitations in availability of resources as well as possible inadequacies in search strategy may have impacted the information obtained and discussed (24).

In conclusion, this report identified pronounced state-level disparities in naltrexone prescribing to Medicaid patients. States that had not expanded Medicaid accounted for nine of the ten lowest prescribing states. Prescribing also decreased during the first quarter of the COVID-19 pandemic.

## Supporting information

Disparities

## Data Availability

All data produced in the present study are available upon reasonable request to the authors

https://data.medicaid.gov/

## Abbreviations

AUD: Alcohol Use Disorder
OUD: Opioid Use Disorder

## References

1. Silva MJ, Kelly Z. The escalation of the opioid epidemic due to COVID-19 and resulting lessons about treatment alternatives. Am J Manag Care. 2020 1;26(7):e202–4.

2. Hoffman J. Soaring overdose rates in the pandemic reflected widening racial disparities [Internet]. The New York Times. 2022 [cited 2022Sep1]. Available from: https://nyti.ms/3v0LBZX

3. Blewett LA, Hest R. Emergency flexibility for states to increase and maintain Medicaid eligibility for LTSS under COVID-19. Journal of Aging & Social Policy. 2020 3;32(4-5):343–9.

4. CDC. Wide-ranging online data for epidemiologic research (WONDER). Atlanta, GA: CDC, National Center for Health Statistics; 2020. Available at http://wonder.cdc.gov.

5. Pollard MS, Tucker JS, Green HD. Changes in adult alcohol use and consequences during the COVID-19 pandemic in the US. JAMA Netw Open. 2020;3(9):e2022942

6. Murthy P, Narasimha VL. Effects of the COVID-19 pandemic and lockdown on alcohol use disorders and complications. Curr Opin Psychiatry. 2021 Jul 1;34(4):376–385.

7. White AM, Castle IP, Powell PA, Hingson RW, Koob GF. Alcohol-related deaths during the COVID-19 pandemic. JAMA. 2022;327(17):1704–6.

8. Pashmineh Azar AR, Cruz-Mullane A, Podd JC, Lam WS, Kaleem SH, Lockard LB, et al. Rise and regional disparities in buprenorphine utilization in the United States. Pharmacoepidemiol Drug Saf. 2020;29(6):708–15.

9. Abraham AJ, Andrews CM, Harris SJ, Friedmann PD. Availability of medications for the treatment of alcohol and opioid use disorder in the USA. Neurotherapeutics. 2020;17(1):55–69.

10. Graves JA, Hatfield LA, Blot W, Keating NL, McWilliams JM. Medicaid expansion slowed rates of health decline for low-income adults in southern states: An analysis of the impact of Medicaid expansion on the self-reported health of low-income older nonelderly adults living in the South. Health Affairs. 2020 1;39(1):67–76.

11. Trivedi MH, Walker R, Ling W, dela Cruz A, Sharma G, Carmody T, Ghitza UE, Wahle A, Kim M, Shores-Wilson K, Sparenborg S. Bupropion and naltrexone in methamphetamine use disorder. N Eng J Med. 2021 14;384(2):140–53.

12. Center for Substance Abuse Treatment. Incorporating Alcohol Pharmacotherapies into Medical Practice. Substance Abuse and Mental Health Services Administration (US); 2009. Treatment Improvement Protocol. Series, No. 49.) Chapter 4—Oral Naltrexone.

13. Bradley KA, Jonas DE. Study examines effectiveness of medications to treat alcohol use disorders - for the media - jama network [Internet]. For The Media JAMA Network. [cited 2022Sep1]. Available from: https://media.jamanetwork.com/news-item/study-examines-effectiveness-of-medications-to-treat-alcohol-use-disorders/#:~:text=The%20NNT%20to%20prevent%20return,(50%20mg%2Fd

14. Oncken C, Van Kirk J, Kranzler HR. Adverse effects of oral naltrexone: analysis of data from two clinical trials. Psychopharmacology (Berl). 2001;154(4):397–402.

15. de Laat B, Nabulsi N, Huang Y, O’Malley SS, Froehlich JC, Morris ED, Krishnan-Sarin S. Occupancy of the kappa opioid receptor by naltrexone predicts reduction in drinking and craving. Mol Psychiatry. 2021; 26(9):5053–60.

16. MACPAC (Medicaid and CHIP Payment and Access Commission). Medicaid and the Opioid Epidemic. 2017.

17. Srivastava AB, Gold MS. Naltrexone: A history and future directions. InCerebrum: the Dana forum on brain science 2018 Sep (Vol. 2018). Dana Foundation.

18. Alinsky RH, Zima BT, Rodean J, Matson PA, Larochelle MR, Adger H, et al. Receipt of addiction treatment after opioid overdose among Medicaid-enrolled adolescents and young adults. JAMA Pediatr. 2020;174(3):e195183.

19. Abraham AJ, Yarbrough CR, Harris SJ, Adams GB, Andrews CM. Medicaid expansion and availability of opioid medications in the specialty substance use disorder treatment system. Psychiatric Serv. 2021;72(2):148–55.

20. CDC. Museum COVID-19 Timeline [Internet]. Centers for Disease Control and Prevention. 2021 [cited 24 November 2021]. Available from: https://www.cdc.gov/museum/timeline/covid19.html#:~:text=January%2020%2C%202020%20CDC,18%20in%20Washington%20state.

21. Corallo B, Moreno S. Analysis of recent national trends in Medicaid and CHIP enrollment [Internet]. Kaiser Family Foundation. 2022 [cited 11 March 2022]. Available from: https://www.kff.org/coronavirus-covid-19/issue-brief/analysis-of-recent-national-trends-in-medicaid-and-chip-enrollment/

22. Ellison A. States ranked by COVID-19 restrictions [Internet]. Becker’s Hospital Review. 2022 [cited 11 March 2022]. Available from: https://www.beckershospitalreview.com/rankings-and-ratings/states-ranked-by-covid-19-restrictions-040821.html

23. Knopf A. Drug decriminalization measure in Oregon would not increase treatment funding. Alcoholism & Drug Abuse Weekly. 2020;32(46):1–3.

24. Liu S, Liu J. Public attitudes toward COVID-19 vaccines on English-language Twitter: A sentiment analysis. Vaccine. 2021;39(39):5499–505.

25. Trofimovitch D, Baumrucker SJ. Pharmacology update: low-dose naltrexone as a possible nonopioid modality for some chronic, nonmalignant pain syndromes. Am J Hosp Palliat Care. 2019;36(10):907–12.

26. Crystal S, Akincigil A, Bilder S, Walkup JT. Studying prescription drug use and outcomes with Medicaid claims data strengths, limitations, and strategies. Med Care. 2007; 45(10 SUPL):S58.

